# Definition of viroIogical endpoints improving the design of HIV cure strategies using analytical antiretroviral treatment interruption

**DOI:** 10.1101/2024.01.26.24301813

**Authors:** Marie Alexandre, Mélanie Prague, Edouard Lhomme, Jean-Daniel Lelievre, Linda Wittkop, Laura Richert, Yves Lévy, Rodolphe Thiébaut

## Abstract

**Background:** Analytical treatment interruption (ATI) is the gold standard in HIV research to validate the ability of novel therapeutic strategies to long-term control viremia without antiretroviral treatment (ART). Viral setpoint is commonly used as endpoint to evaluate their efficacy. However, to mitigate higher viremia risk without ART, trials use short ATI phases and strict virological ART re-start criteria, compromising the observation of the setpoint.

**Methods:** We analyzed viral dynamics in 235 HIV-infected participants from three trials, examining various virological criteria during ATI phases. Time-related (e.g. time to rebound, peak and setpoint) and VL magnitude-related criteria (peak, setpoint and time-averaged AUC [nAUC]) were described. Spearman correlations were analyzed to identify surrogate endpoints for setpoints. Additional correlation analyzes were performed to identify optimal virological ART re-start criteria mitigating the risks of ART interruption and the evaluation of viral control.

**Results:** Comparison of virological criteria between trials showed strong dependencies on ATI design. Similar correlations were found across trials, with nAUC identified as the criterion most strongly correlated with the setpoint, with correlations higher than 0.70. A threshold of at least 100,000 copies/mL for two consecutive VL measurements is requested as virological ART re-start criteria to keep strong correlations between the setpoint and nAUC.

**Conclusions:** Our results emphasize the benefits of an ATI phase longer than 12 weeks, with regular monitoring, and a VL threshold of 100,000 copies/mL as virological ART re-start criteria to limit the risk for patients while capturing enough information to keep nAUC as an optimal proxy for the setpoint.

## Introduction

Treatment interruption, i.e., discontinuation of antiretroviral treatment (ART), has been used in the evaluation of strategies aimed to save drug exposure (1–3). However, interruption of ART is associated to a higher risk for clinical progression (4). Nevertheless, treatment interruption remains the only means to evaluate therapeutic interventions such as immunotherapies or therapeutic vaccines (5) aimed to control HIV replication without ART (6–11).

While accepted in a context of clinical trials involving people living with HIV (PLWH) (12,13), treatment interruption should be carefully monitored and several recommendations have been proposed to limit potential risks, in terms of morbidity, mortality, disease progression, HIV transmission, emergence of new drug-resistance, development of neurological or cardiovascular disorders (14,15). These recommendations led to the development of clearly- defined interruptions: Analytical treatment interruptions (ATI) (14). Therefore, it is of the upmost importance to optimize the design of ATI trials to provide a maximum information with a minimum risk exposure (16). Stringent ART re-start criteria upon viral rebound are included in research protocols while virological endpoints to assess efficacy of experimental strategies have been largely discussed (14,16–20). Two types of criteria are usually considered in the definition of virological endpoints, each leading to a distinct design of the ATI phase: the time to viral rebound and the setpoint, that is the relatively stable level of HIV viral load (VL) after the rebound (14,21). When the outcome of interest is the time to viral rebound, the threshold of VL for ART re-start is often set at a low level to limit risks (e.g. 1 000 copies/mL). Optimal designs have been recently proposed for studies using “time to viral rebound” as endpoints (16). One drawback here could be the ART re-start at a low viral rebound which impair the evaluation of viral replication control. On the other hand, studies using “setpoint” as endpoint require longer treatment interruption periods with higher thresholds for ART re-start, leading to a potential increased risk of HIV transmission and side effects. Thus, while the “time to rebound” could be easily determined in ATI studies, assessing the setpoint of viral replication, a proxy of the control of viral rebound, remains challenging. Therefore, the need to define optimal designs of ATI phase for “setpoint” studies, mitigating the risks related to treatment interruption and the evaluation of viral control, is essential.

In the present study, we analyzed several virological parameters of the kinetics of viral rebound following ATI in various clinical trials of immunotherapeutics in chronic PLWH. The objective was to identify at earliest phase of ATI potential surrogate endpoints predictive of viral setpoints. These results might have a practical impact on the design of future clinical trials with ATI.

## Materials and Methods

### Clinical trial data

We considered individual data extracted from three distinct trials in PLWH including two therapeutic vaccine trials and one testing an IL-2 immunotherapeutic strategy. All trials comprised an ATI phase to evaluate the virological efficacy of these strategies in maintaining low viral replication after ART interruption (**Figure 1**).

**Figure 1.**
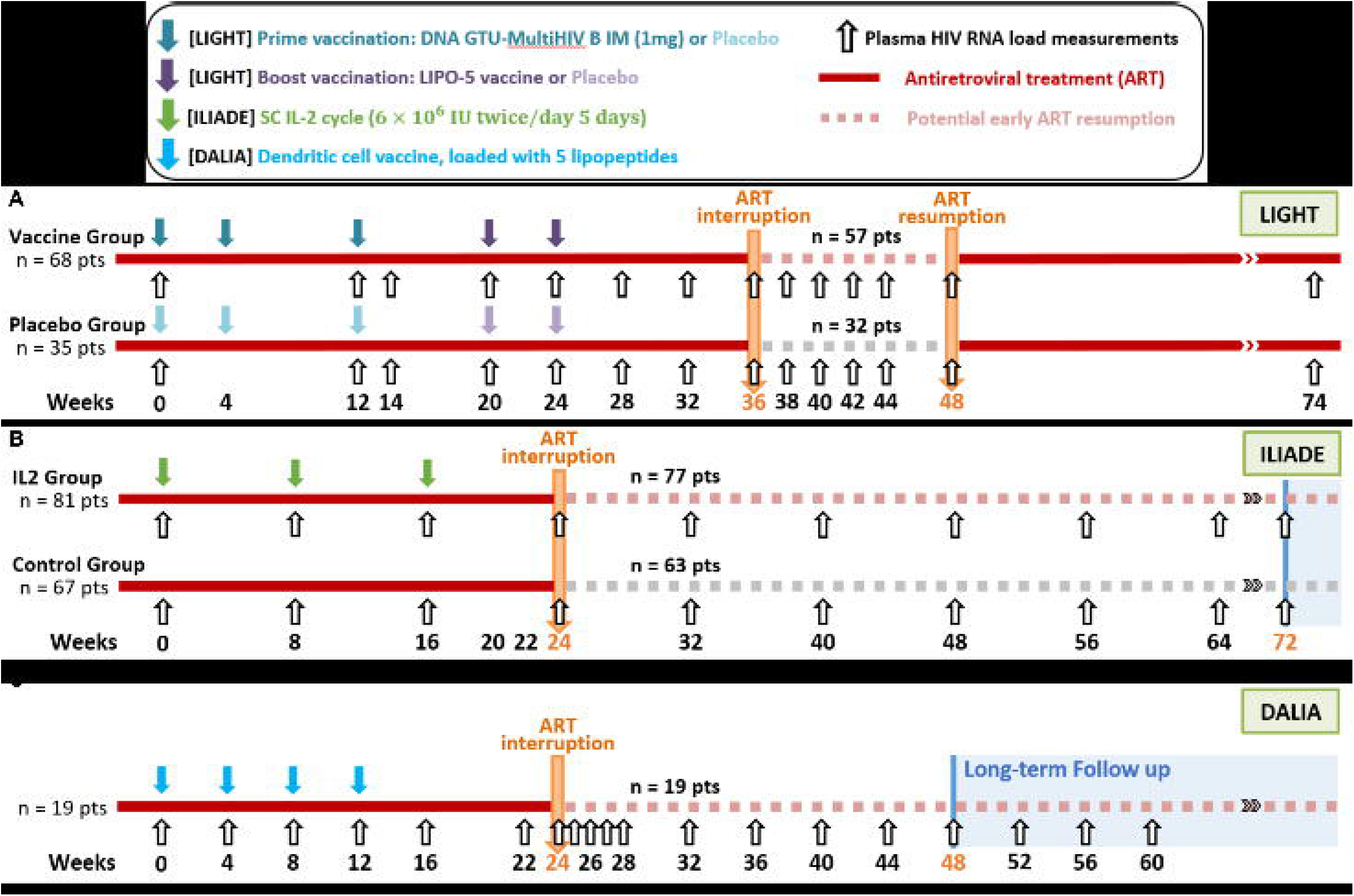
Study designs of HIV therapeutic vaccine trials. (A) L IGHT trial. Participants received a prime-boost vaccine strategy, using recombinant DNA vaccine GTU-MultiHIV B or placebo as prime (3 doses at weeks 0, 4 and 12), and lipopeptide vaccine HIV-LIPO-5 or placebo as boost (2 doses at weeks 20 and 24). (B) ILIADE trial. Participants an IL-2 immunotherapy, defined by three cycles of subcutaneous injections of recombinant IL-2 (2 injections of 6x106 international 395 units (IU) per day for 5 days, at weeks 0, 8 and 16), or placebo. (C) DALIA trial. Participants received 4 doses of a dendritic cell vaccine, at weeks 0, 4, 8 and 12 weeks. (A-C) In the three trials, patient could prematurely restart ART during the protocol-defined ATI phase if they met the following criteria: (1) wish of patients or of their doctors, (2) CD4+ T-cell count dropped below 350 cells/μL at two consecutive measurements 2-weeks apart, and (3) occurrence of an opportunistic infection or a serious non-AIDS defining event. In DALIA, a drop of CD4+ T-cell count below 25% of total lymphocytes for two consecutive measurements was also considered. Analysis performed in this paper focused exclusively on viral load data collected during ATI phase (i.e., free-of-ART phase). In ILIADE and DALIA, only data collected from ART interruption to the time of primary endpoint (colored in orange, W72 and W48, respectively) were considered.

The VRI02 ANRS 149 LIGHT trial (22) (NCT01492985, and labelled LIGHT) was a multicenter, double-blind, placebo-controlled, randomized, Phase II trial evaluating the safety, immunogenicity and virological efficacy of a prime-boost vaccine strategy, using recombinant DNA vaccine GTU-MultiHIV B as prime, and lipopeptide vaccine HIV-LIPO-5 as boost.

The ANRS/VRI DALIA trial (23–25) (NCT00796770, and labelled DALIA) was a single-center, single arm Phase I trial evaluating the safety of a dendritic cell vaccine.

The ANRS 118 ILIADE trial (26) (NCT00071890, and labelled ILIADE) was a multicenter, open- label, placebo-controlled, randomized Phase II/III trial evaluating the efficacy of IL-2 immunotherapy to extend the period of time off ART, defined by three cycles of subcutaneous injections of recombinant IL-In LIGHT, 103 ART-treated HIV-infected patients were randomized in a 2:1 ratio to receive their ART in combination with either an active vaccine or placebo. From week 36 to 48, patients interrupted their ART and were regularly monitored with immunological (CD4+ and CD8+ T cell counts) and VL measurements performed every 2 weeks (**Figure 1A**). The primary virological endpoint studied was the maximum observed VL (in log_10_ copies/mL) during ATI phase.

In DALIA, 19 ART-treated HIV-infected patients were included who interrupted ART for 24 weeks, from week 24 to 48, and were monitored every 1 to 4 weeks (**Figure 1C** ). No virological endpoint was evaluated in this study.

Finally, ILIADE enrolled 148 ART-treated HIV-infected patients randomized in a 1:1 ratio to receive ART either in combination with IL-2 therapy or alone. From week 24 to 72, ART was interrupted and CD4+, CD8+T cells counts and VL were collected every 8 weeks (**Figure 1B**). No virological endpoint was considered in ILIADE.

The reader can refer to **Figure 1** for the list of criteria leading to ART re-start before the end of the protocol-defined ATI phase.

### Ethics statement

The protocol of LIGHT study was approved by the ethics committee of Ile de France 5 (Paris- Saint-Antoine) and authorized by the French regulatory authority (ANSM). The DALIA study was sponsored by the Baylor Institute for Immunology Research and the Agence Nationale de Recherches sur le SIDA et les hepatites (INSERM ANRS), and the protocol was approved by the IRB of Baylor Research Institute (BRI). The protocol of ILIADE study was approved by the ethics committee of the Hospital Henri Mondor and the National Institute of Allergy and Infectious Diseases (NIAID). All participants provided written informed consent before participation.

### Definition of virological criteria

Virological data of the trials described above were analyzed in the present study, and a total of eight criteria summarizing the dynamics of viral rebound during ATI were considered. To account for the differences of design of ATI phases between the three trials (e.g., ATI phase duration or sampling frequencies, virological criteria were divided into two categories: (1) fixed criteria if they were principally related to the magnitude of VL and, (2) temporal criteria if they corresponded to time-related outcomes.

The three fixed criteria included in this analysis were (1) peak VL, (2) setpoint and, (3) time-averaged area under the curve (AUC) (i.e., AUC divided by the duration of ATI phase, labelled nAUC). A normalized version of AUC was used to adjust for ATI duration and to be comparable between patients. In the absence of a formal definition of the setpoint in the literature, the steady state of VL reached after the peak VL was defined as the mean of observations included in a sliding window of height 0.5 log_10_ copies/mL, with a width of at least 4 weeks and including at least two VL values (17). Moreover, the assumption that the setpoint was correctly measurable in the different trials was made.

The temporal criteria were (1) time to rebound (TTR), (2) slope of viral rebound, (3) time to peak VL, (4) time to setpoint, (5) time to distinct VL thresholds (i.e., 200, 1 000, 10 000 and 100 000 copies/mL). The reader can refer to **Table 1** and **Figure 2A** for a full definition of these criteria

**Figure 2.**
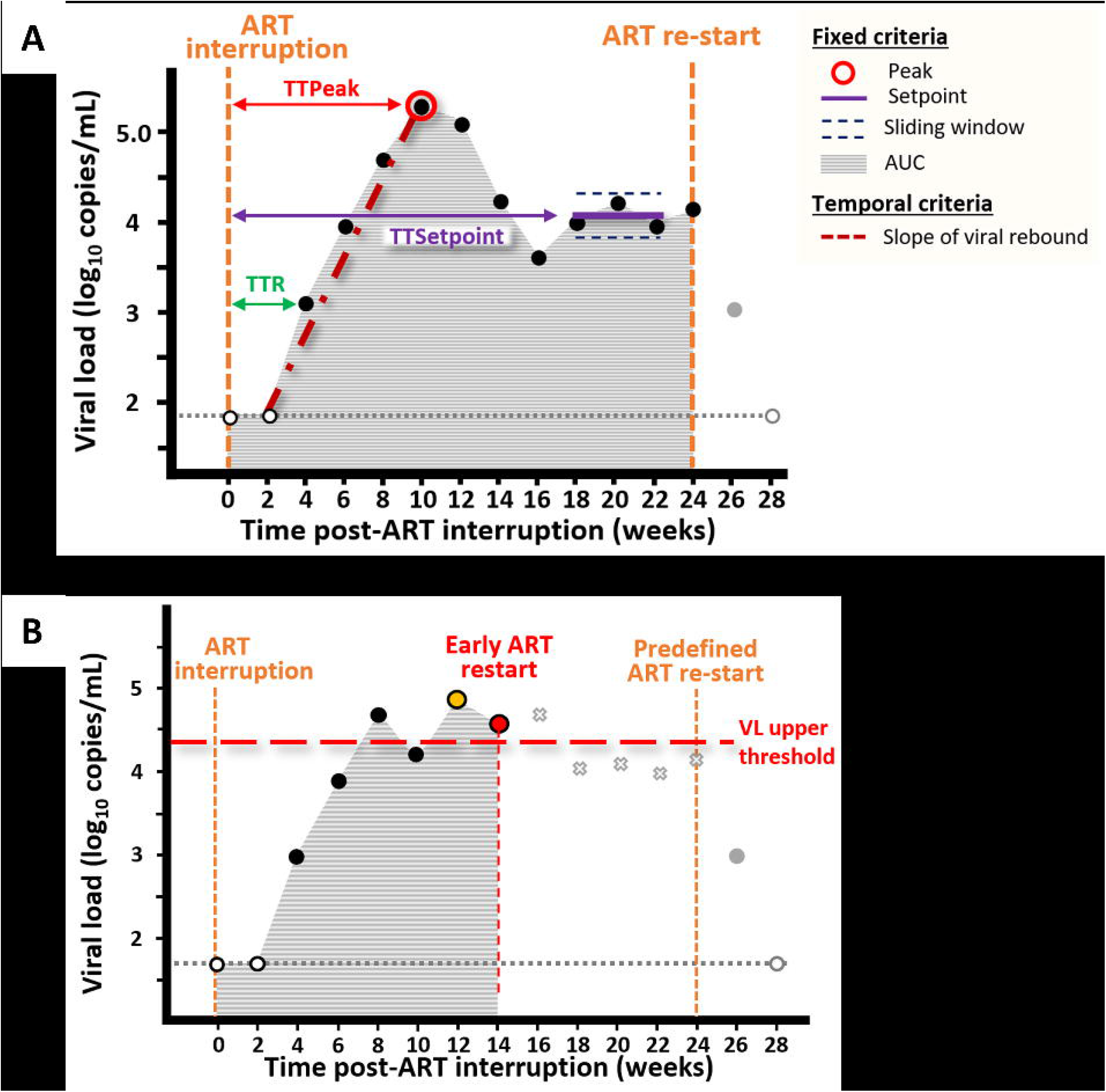
Graphical representation of virological criteria analyzed in the study. Black and white dots represent detectable and undetectable viral load measurement collected during ATI phase, respectively, and gray dots those collected after ART re-start. The horizontal gray dotted line represent the limit of detection. **(A)** Virological criteria describing viral rebound dynamics. **(B)** Virological restart ART criteria inducing strategy discontinuation and censoring of follow-up. Orange dot shows the first viral load measure exceeding the VL upper-threshold represented by the horizontal red dashed line (i.e., time at which viral dynamic is censored when no confirmation measure is considered before ART re-start). Red dot shows the viral load measure at which viral dynamic is censored when a confirmation measure is required before any ART resumption. Gray crosses display viral load measures unobserved due to censoring of following after strategy discontinuation. Abbreviations: ART, antiretroviral treatment; TTR, time to rebound; TTsetpoint, time to setpoint; TTPeak, time to VL Peak; AUC, area under the curve; VL, viral load.

**Table 1.** Definitions of virological criteria analyzed in the study.

| **Virological criteria** | **Definition** |
| --- | --- |
| **Fixed criteria** |  |
| **Peak VL**<br>*[in log<sub>10</sub> copies/mL]* | The maximum VL value during ATI phase.<br>In case of long ATI phase (e.g., in ILIADE trial), an increasing viral dynamics can be observed in some patients after the stabilization of the dynamics and reach higher values than the peak of rebound. In that case, the higher VL value observed before the stabilization was kept. |
| **Setpoint**<br>*[in log<sub>10</sub> copies/mL]* | The mean value of VL values belonging to the first sliding window with a height of 0.5 log<sub>10</sub> copies/mL, and with a width of at least 4 weeks including at least two consecutive VL values. |
| **Time-averaged AUC (nAUC)**<br>*[in log<sub>10</sub> copies/mL]* | Area under the viral rebound curve averaged by the duration (in weeks) of ATI phase. |
| **Temporal criteria** |  |
| **Time to rebound (TTR)**<br>*[in weeks]* | The time between ART interruption and the first detectable VL value (>40 copies/mL in LIGHT, and >50 copies/mL in ILIADE and DALIA). |
| **Slope of viral rebound**<br>*[in log<sub>10</sub> copies/mL/week]* | The slope from the last undetectable VL value to the peak VL. |
| **Time to peak VL**<br>*[in weeks]* | The time between ART interruption and the peak VL (only for patients with observed peak VL). |
| **Time to setpoint**<br>*[in weeks]* | The time between ART interruption and the first VL value belonging to the window defining the setpoint (only for patients with observed setpoint). |
| **Time to VL threshold**<br>*[in weeks]* | The time between ART interruption and the first VL value exceeding a specific VL threshold.<br>Four different VL thresholds, commonly used in HIV vaccine trials, were considered: 200, 1 000, 10 000, and 100 000 copies/mL. |

Abbreviations: ATI, Analytical treatment interruption; VL, Viral load; ART, Antiretroviral treatment.

### Identification of the optimal virological criterion to be used as primary endpoint

We sought to identify optimal criterion summarizing viral dynamics that could be used as proxy for the setpoint, based on the following characteristics: highly associated with the setpoint, and whose observation is less impacted by the duration of ATI phase.

Individual viral dynamics can be impacted by both the effect of the intervention and the design of ATI phase. In each trial, the design of ATI being comparable between the two groups of treatment, we first performed a descriptive analysis and a comparison of the two groups at each time point to identify potential differences of dynamics, induced by treatment effect, that could impact the values of virological criteria. As no statistical differences were identified between groups, whether in LIGHT or ILIADE and whatever the time point, the two groups were pooled together in each trial to increase statistical power. The following analyses were then performed at trial level (i.e., LIGHT Vs ILIADE Vs DALIA). Secondly, once the eleven aforementioned virological criteria estimated for each individual, a descriptive and comparison analysis between trials were performed to analyze how ATI designs impacted their values. Finally, Spearman correlations were evaluated between criteria and compared between trials to (1) see how ATI designs impact correlation patterns, and (2) identify criterion the most associated to the setpoint.

In this analysis, patients were excluded if they fulfilled one of the following criteria: ATI not initiated, detectable VL at the time of ART interruption (condition for ATI initiation), therapeutic strategy not received (e.g., at least one IL-2 dose requested in ILIADE). Moreover, only the first 48 weeks of ATI (i.e., until the primary outcome at week 72, see Figure 1B) were included in ILIADE, to limit heterogeneity among trials.

### Assessment of the choice of virological restart ART criteria on the evaluation of the virological endpoint

To limit the risks for participants during ATI phase, ART re-start criteria are defined in protocols. These criteria define conditions under which it is vital for a patient to re-start ART before the end of ATI phase. While the patient remains in the trial and is still frequently monitored after ART re-start to ensure its safety and to contribute to other endpoints, this one does not fulfill conditions to contribute to virological endpoint. Consequently, censoring of follow-up is considered due to strategy discontinuation and virological measurements collected from that point are treated as monotonic missingness (once missing, always missing subsequently) for the evaluation of virological criteria. Although ART re-start criteria can have a strong impact on primary outcomes and, by extension, on the evaluation of vaccine efficacy, no real consensus has been reached about their definition (14). In this work, we focused on virological ART re-start criteria defined as VL threshold above which patients have to re-start ART for their safety, named VL upper-threshold thereafter. As shown in **Figure 2B** , censored follow-up, resulting from the use of VL upper-threshold, required the study of right-censored viral dynamics.

We re-analyzed the three trials by investigating the impact of VL upper-thresholds on the association between the setpoint and nAUC. Three commonly considered VL upper- thresholds were investigated: (1) 10 000 copies/mL, (2) 50 000 copies/mL, and (3) 100 000 copies/mL. To this end, Spearman correlations were studied between the setpoint, estimated on complete data (i.e., not impacted by VL upper-threshold), and the nAUC calculated on right-censored viral dynamics, labelled right-censored nAUC (**Figure 2B** ). Moreover, we investigated the need for a confirmation measure before any ART restart. To this end, correlations between the setpoint and the nAUC were evaluated considering censoring of follow-up either 1) as soon as VL upper-threshold has been exceeded for one viral measure (i.e., without confirmation measure), or 2) once VL upper-threshold has been exceed for two consecutive viral measures (i.e., with confirmation measure).

### Statistical analysis

All analyses were performed in R (version 4.2.1). Continuous variables were expressed in median (interquartile [IQR]), and discrete variables in absolute number and percentage. The Student t-test was used to compare VL measurements between groups. The non-parametric Mann-Whitney U and Kruskal-Wallis tests were used to perform pairwise comparisons and multiple group comparisons of virological criteria, respectively. The associations between virological criteria in each trial were evaluated using Spearman correlations. Statistical significance was defined as a p-value lower than 0.05, after Benjamini and Hochberg false discovery rate-adjustment (27) for multiple testing.

## Results

### Population

A total of 235 patients, out of the 270 enrolled in the three trials, were included in the present study: 87 in LIGHT (n=31 in placebo group and n=56 in vaccine group), 129 in ILIADE (n=57 in control group and n=72 in IL-2 group) and 19 in DALIA **(Figure S1).**

### Description of virological measurements

Viral rebound dynamics for the three trials are shown in **Figure 3** , and distributions of VL measured at each sampling time point during ATI phase are summarized in **Table S1**. The median length of follow-up before ART re-start ranged from 12 to 48 weeks, with a median number of VL measurements from 6 to 11 (**Table 2**). The percentages of patients prematurely reinitiating their ART before the end of ATI phase were equal to 21 % in LIGHT (23% for Placebo and 20% for vaccinated), 26% in ILIADE (35% for Control and 19% for IL-2), and 16% in DALIA.

**Figure 3.**
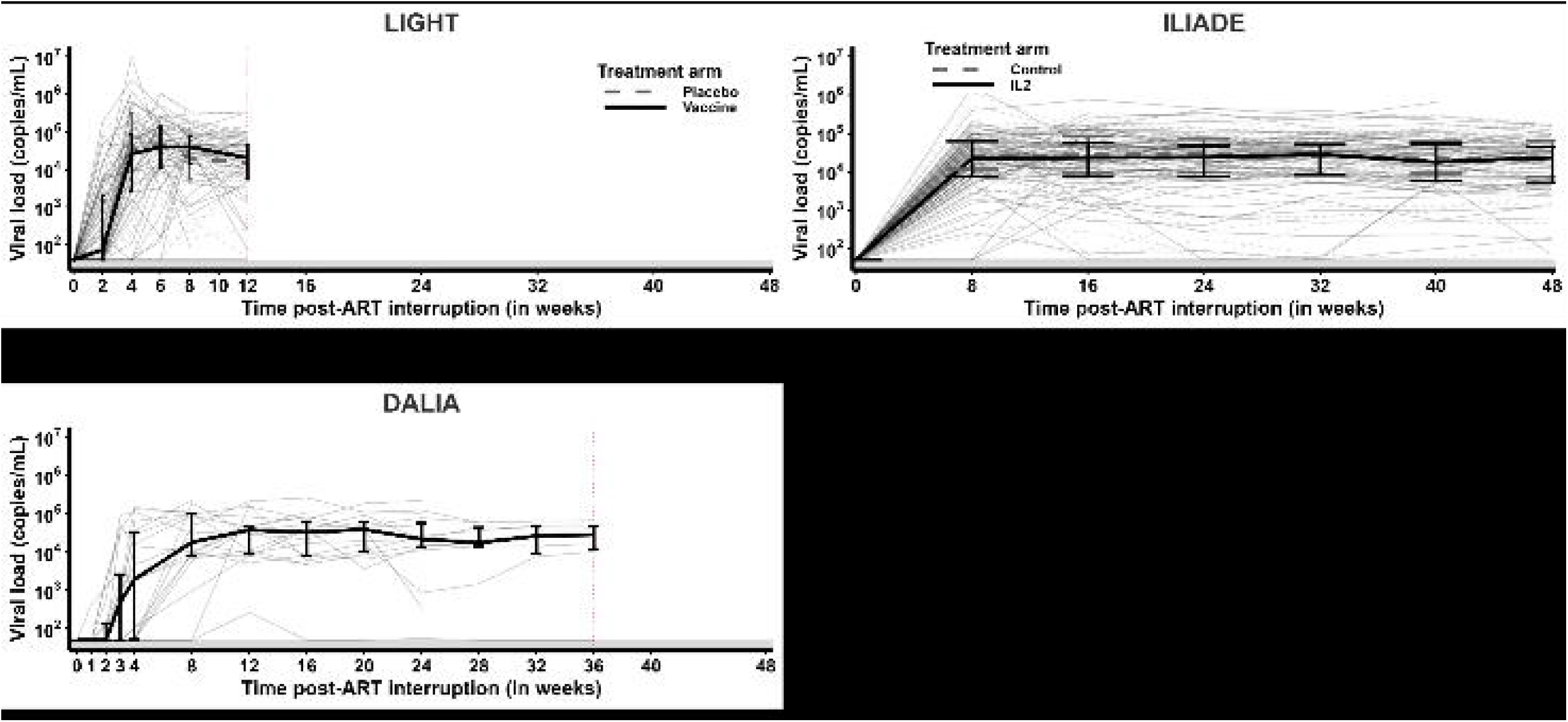
Viral dynamics observed in the three trials in ATI phase. (A) Viral dynamics observed during 12-week ATI phase in LIGHT trial, with monitoring every 2 weeks. **(B)** Viral dynamics observed during 48-week ATI period in ILIADE trial, with monitoring every 8 weeks. **(C)** Viral dynamics observed during 36-week ATI period in DALIA trial, with monitoring every 1 to 4 weeks. **(A-C)** Thick and thin lines represent the median and individual dynamics, respectively, and error bars represent interquartile ranges. Solid and dashed lines correspond to dynamics in therapeutic and control/placebo groups, respectively. Grey areas highlight limits of detection (i.e., 40 cp/mL for LIGHT, and 50 cp/mL for ILIADE and DALIA). Red dotted vertical lines for represents the end of ATI phase (week 12 in LIGHT and week 36 in DALIA).

**Table 2.** Description of ATI in the three therapeutic vaccine trials.

|  | LIGHT (n=87) |  | ILIADE (n=129) |  | DALIA (n=19) |
| --- | --- | --- | --- | --- | --- |
|  | Placebo (n=31) | Vaccine (n=56) | Control (n=57) | IL2 (n=72) |  |
| **ATI duration (weeks)** |  |  |  |  |  |
| Median [Min-Max] | 12 [4 ; 12] | 12 [2 ; 12] | 48 [0 ; 48] | 48 [0 ; 48] | 28 [8 ; 36] |
| **Nb VL measurements** |  |  |  |  |  |
| Median [Min-Max] | 6 [3 ; 6] | 6 [2 ; 6] | 7 [1 ; 7] | 7 [2 ; 7] | 11 [6 ; 13] |
| **ATI re-start \*** |  |  |  |  |  |
| n (%) | 7 (23) | 11 (20) | 20 (35) | 14 (19) | 3 (16) |

Abbreviations: ATI, Analytical treatment interruption; VL, Viral load; ART, Antiretroviral treatment.
\*Evaluation of the number and the percentage of patient who restart their ART before the end of ATI phase, i.e. before predefined ATI duration in LIGHT (12<sup>th</sup> week) or before time of primary endpoint in ILIADE (48<sup>th</sup> week) and DALIA (24<sup>th</sup> week).

Comparisons, at the different time of measurements, between VL values collected in the experimental and the control group highlighted similar viral, whether in LIGHT (p-values > 0.43) or in ILIADE (p-values > 0.53) (see **Table S1**). Groups were then pooled in each trial to increase the statistical power. As shown in **Figure 4**, no significant difference was observed between the different trials from week 8 (p-value > 0.28). However, viral dynamics differed across trials during the earliest phase of ATI. Mean VL measured in DALIA were significantly lower than in LIGHT at weeks 2 and 4 (2.05 vs 2.49 log10 copies/mL at week 2, p-value=0.04; 3.26 vs 4.12 log_10_ copies/mL at week 4, p-value=0.04).

**Figure 4.**
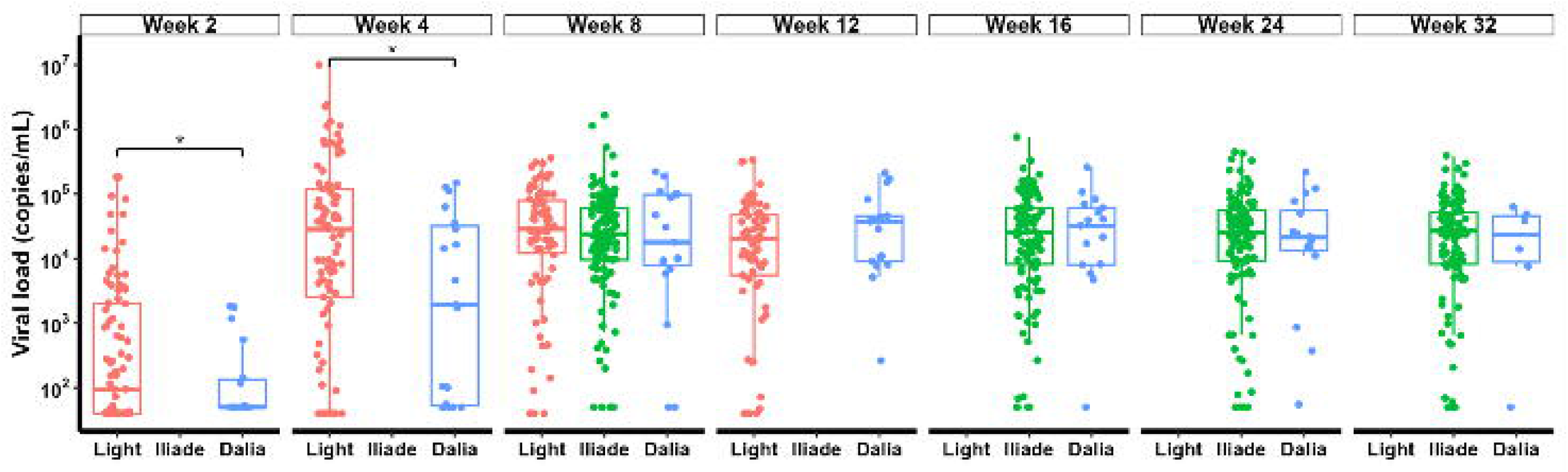
Distribution of viral load measurements collected during ATI phase in the three trials at common time points. (LIGHT in blue, ILIADE in green, and DALIA in pink). Brackets show significant p- values after multiplicity adjustment: *: p ≤ 0.05, **: p ≤ 0.01, ***: p ≤ 0.001, ****: p ≤ 0.0001.

Despite the absence of strong differences in the dynamics of viral rebound between trials, the duration of ATI phase, sampling frequencies and intervals differ between trials, and consequently can impact values of virological criteria.

### Assessment of different virological criteria definitions

The virological criteria describing dynamics of viral rebound calculated in the different groups are summarized in **Table 3**. No statistical differences were found between groups in LIGHT (Placebo vs Vaccine) and ILIADE (Control vs IL-2), allowing an analysis at trial level in the following results (**Figure 5**).

**Figure 5.**
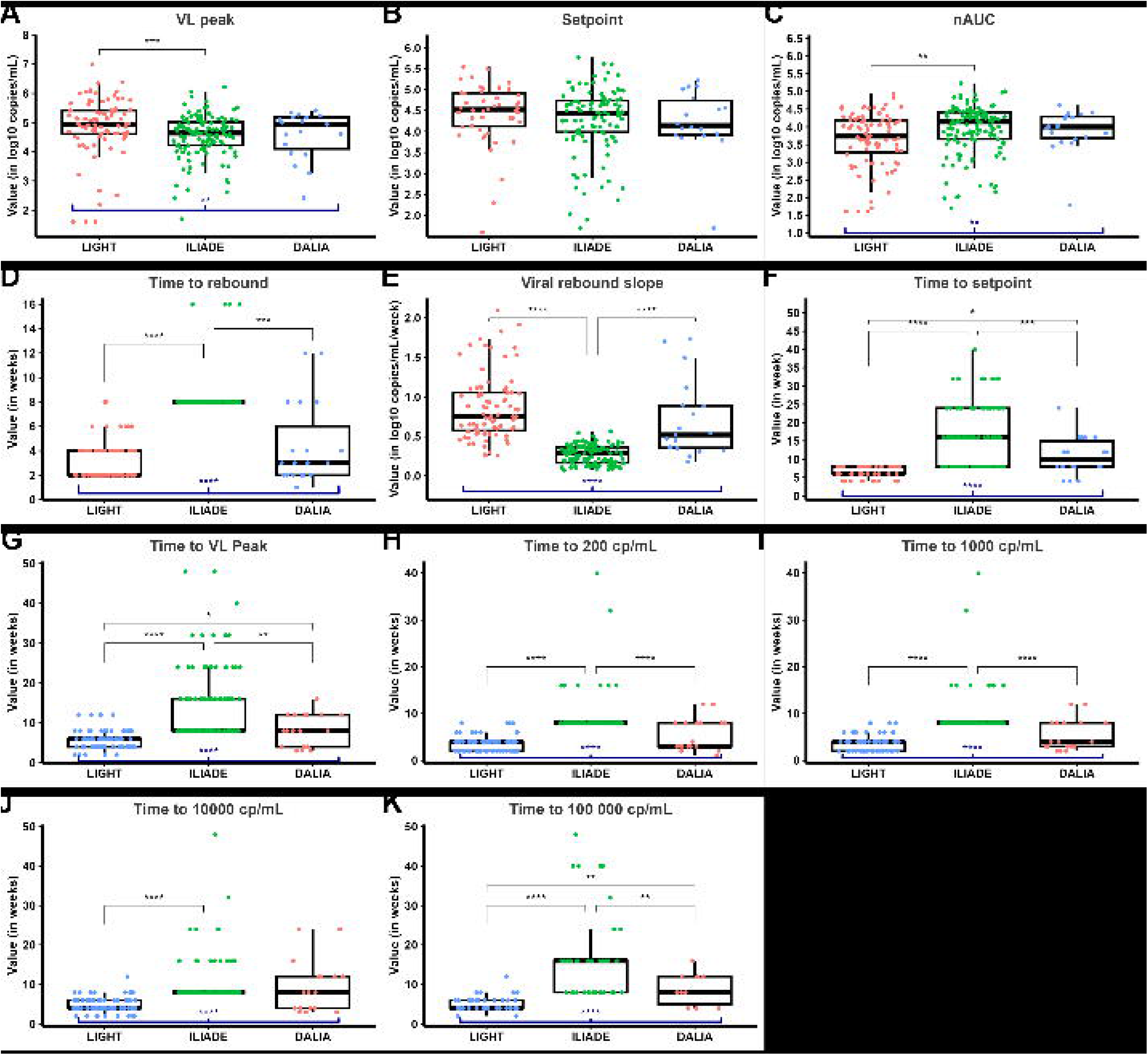
Distribution of different virological criteria within each trial. Blue thick brackets below boxplots and black thin brackets above boxplots show significant p-values of the multiple group comparisons (Kruskal-Wallis test) and pairwise comparisons (Mann-Whitney U test), respectively. Significant p-values after multiplicity adjustment: *: p ≤ 0.05, **: p ≤ 0.01, ***: p ≤ 0.001, ****: p ≤ 0.0001. **(A)** Distribution of the peak VL, in log_10_ cp/mL. **(B)** Distribution of the setpoint, in log_10_ cp/mL. **(C)** Distribution of the time-averaged AUC (nAUC), in log_10_ cp/mL. **(D)** Distribution of the time to rebound, in weeks. **(E)** Distribution of the slope of the viral rebound, in log_10_ cp/mL/week. **(F)** Distribution of the time to setpoint, in weeks. **(G)** Distribution of the time to peak of VL, in weeks. **(H)** Time to 200 copies/mL of VL, in weeks. **(I)** Time to 1 000 copies/mL of VL, in weeks. **(J)** Time to 10 000 copies/mL of VL, in weeks. **(K)** Time to 100 000 copies/mL of VL, in weeks. Abbreviations: nAUC, time-averaged area under the curve ; VL, viral load.

**Table 3.**
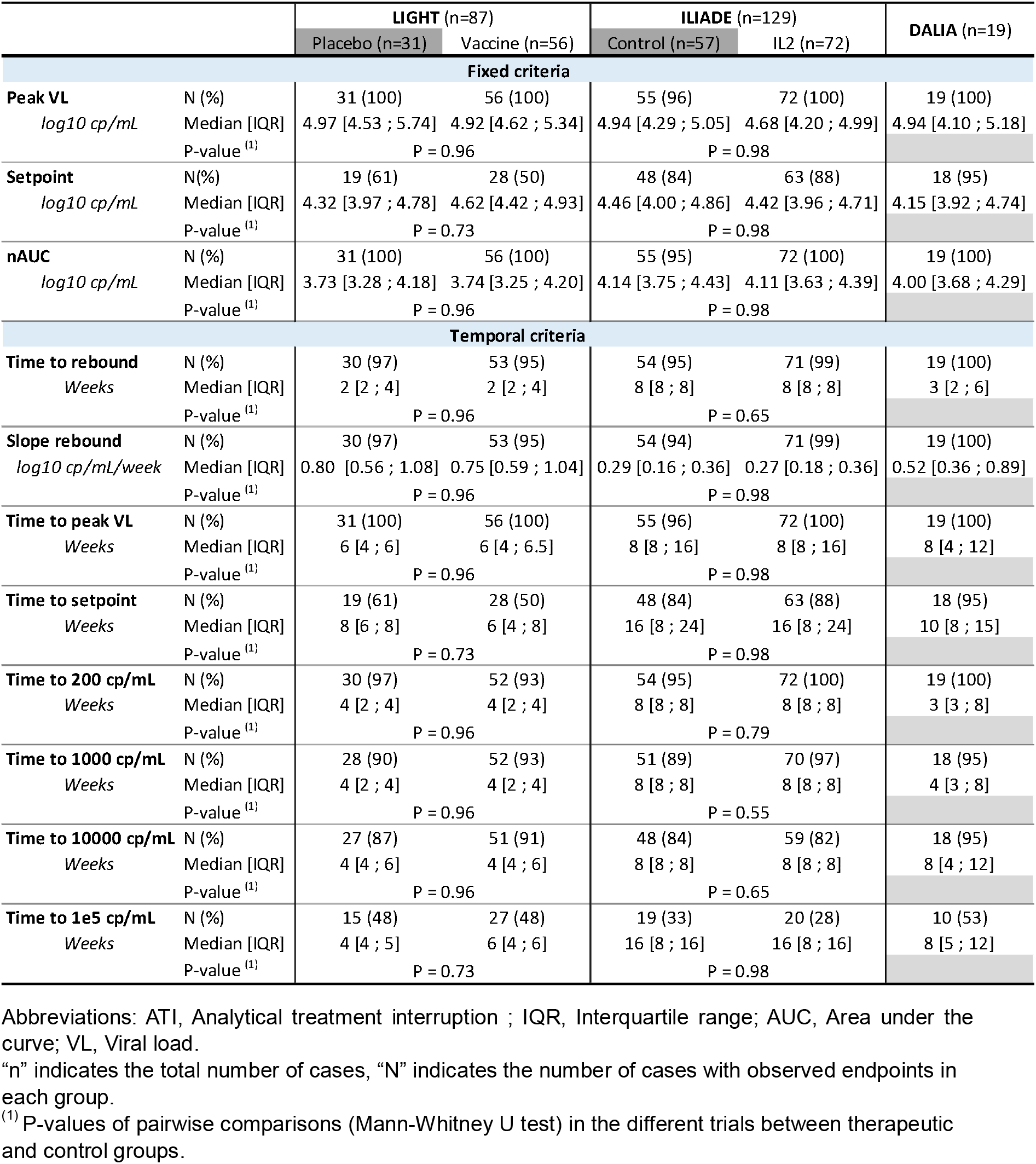
Distribution of the different virological criteria estimated in the three therapeutic vaccine trials.

#### Fixed criteria

No statistical differences of setpoints were observed between trials with mean values ranged from 4.23 in DALIA to 4.42 log_10_ copies/mL in LIGHT (p-value=0.39) (**Figure 5B**). This result was in accordance with the absence of significant differences of VL measurements beyond week 4. Despite this homogeneity among trials, a lower percentage of patients reached the setpoint during ATI in LIGHT (54%) compared to both DALIA (95%) and ILIADE (86%). This difference can be explained by the shorter ATI phase of only 12 weeks in LIGHT, making its observation more difficult.

Both the peak VL and the nAUC were significantly different between LIGHT and ILIADE, whereas no differences were observed between DALIA and LIGHT, DALIA and ILIADE. As shown in **Figure 5A** and **Figure 5C**, the mean peak VL was significantly higher in LIGHT than in ILIADE (+ 6%, p-value <0.001), while the mean nAUC was significantly lower in LIGHT than in ILIADE (- 8%, p-value <0.01). Though not a direct time-dependent criteria, the lower peak VL in ILIADE can be explained by its study design: the longer the time-interval between two VL measurements (every 8 weeks in ILIADE instead of 2 weeks in LIGHT), the higher the probability to miss the peak and capture a measure in the decreasing dynamic following the peak (see **Figure S3** for an illustration).

#### Time-dependent criteria

The analysis of time-dependent criteria highlighted their strong dependence on study design, i.e. ATI duration and sampling intervals. As shown in **Table 3** and **Figure 5 D-F** , with VL measurements collected every 8 weeks, ILIADE’s patients showed significantly different results than those included in LIGHT and DALIA. In particular, they were characterized by a significantly higher mean TTR and time to setpoint (TTR: 8.64 vs 3.04 and 4.47 weeks, respectively, p-values < 0.001 ; time to setpoint: 15.9 vs 6.43 and 10.9 weeks, respectively, p-values <0.001). In contrast, a significantly lower mean slope of rebound was observed in ILIADE than in both LIGHT and DALIA (-69% and -62%, respectively, p-value <0.0001). These variations mostly result from the higher TTR. Except for barely significant lower mean time to setpoint between LIGHT and DALIA (6.43 vs 10.9 weeks, p-value < 0.05), explained by the difference of ATI duration, similar temporal endpoints were observed in these two trials.

### Analysis of correlations between virological criteria

To identify virological criteria summarizing viral rebound and highly associated with the setpoint, we performed correlation analysis between the virological criteria. **Figure 6** displayed Spearman correlations between the five virological criteria the most used obtained in the three trials (see **Figure S4** for correlations between the eleven virological criteria defined in this analysis, and **Figure S5** for Pearson correlations and scatter plots). Similar patterns of correlations were obtained between the different trials. First, the absence of significant correlations between the TTR and the setpoint, the two virological endpoints the most widely recommended to assess vaccine efficacy in HIV therapeutic clinical trials (14). Second, we identified strong significant positive correlations between the setpoint and both the peak VL and the nAUC (p-values < 0.001), with higher correlation coefficients for the nAUC (0.70 vs 0.61 in LIGHT, 0.90 for both in ILIADE, and 0.75 vs 0.73 in DALIA). The correlations between TTR and other endpoints were less consistent across trials. For instance, a negative correlation between the TTR and the nAUC was found in LIGHT (-0.57, p-value <0.0001) but not in DALIA and ILIADE.

**Figure 6.**
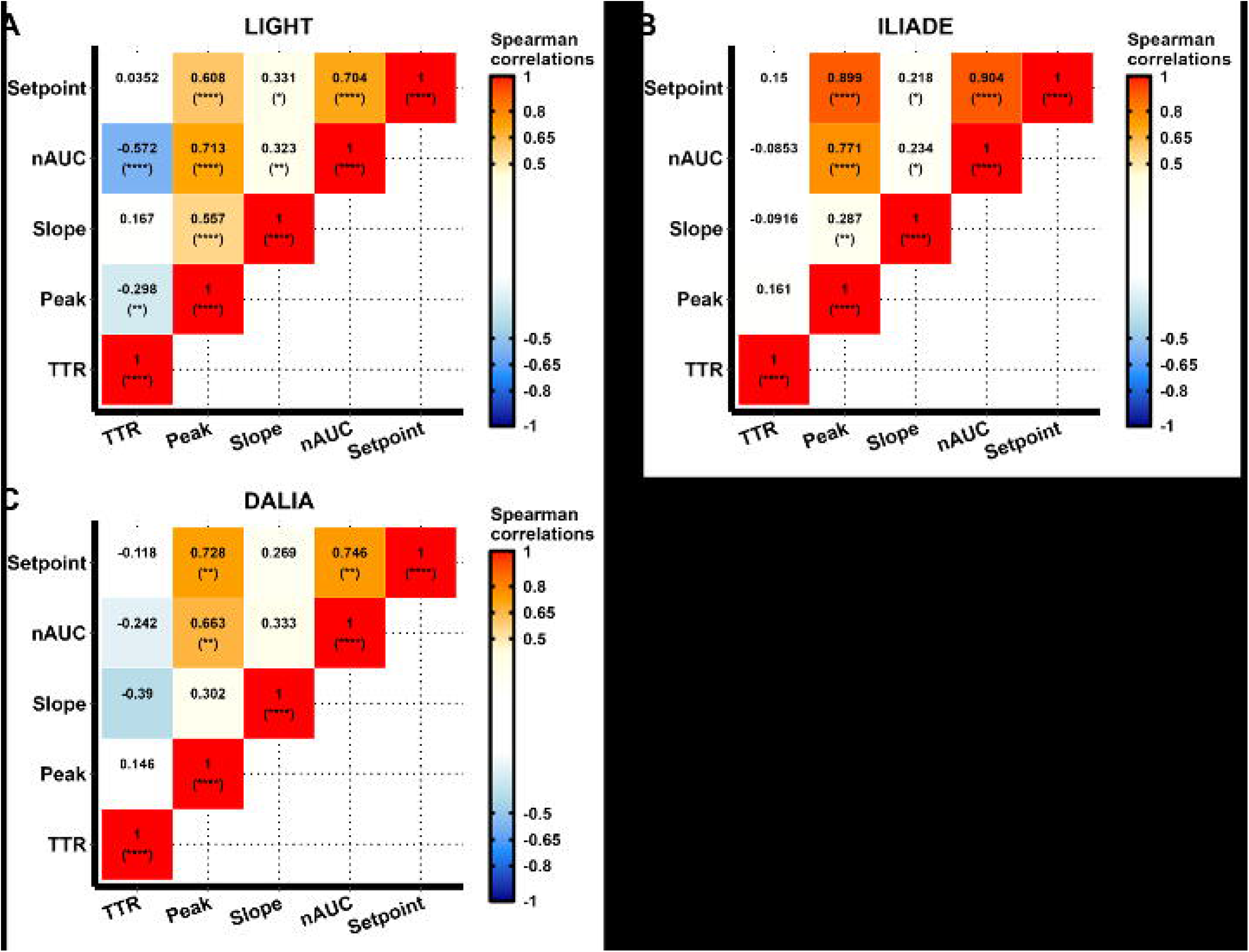
Heatmap of pairwise Spearman correlations between the five main virological criteria in the three clinical studies. (A) LIGHT clinical trial (n=87). **(B)** ILIADE clinical trial (n=129). **(C)** DALIA clinical trial (n=19). **(A-C)** Correlations higher than 0.5 are colored in orange-red palette and correlations lower than -0.5 in blue palette. P-values adjusted for multiplicity testing are indicated in brackets: *: p ≤ 0.05, **: p ≤ 0.01, ***: p ≤ 0.001, ****: p ≤ 0.0001. Abbreviations: nAUC, time-averaged area under the curve; TTR: time to rebound.

These results allowed to identify the nAUC as the virological criteria the most strongly correlated with the setpoint, and which can be evaluated regardless the design of the ATI phase. However, it was worth noting that the strongest correlations between these two criteria were observed in ILIADE rather than in LIGHT, indicating that this association seems influenced by the duration of ATI phase.

### Virological restart criteria impact the association between the setpoint and the nAUC

We assessed the impact of ART re-start criteria, defined as VL upper-threshold (see **Figure 2B**), on the correlation between the setpoint and the nAUC. **Figure 7** illustrated the Spearman correlations between the setpoint and the right-censored nAUC for different values of the VL upper-threshold (whether for discrete values in **Figure 7A** or continuous values in **Figures 7B-D** ), with or without considering a confirmation measure before censoring follow-up. In most cases, the higher the VL upper-threshold, the more significant and stronger the correlation coefficient. Moreover, results obtained in both LIGHT and DALIA highlighted the benefit of considering a confirmation measure to retrieve the significance of correlations. While no significant correlations were identified in these two trials in absence of confirmation measure, positive significant correlations were identified when an additional VL measure was considered with a VL threshold of 100 000 cp/mL. The use of a low threshold to censure data can lead to non-conclusive results (non-significant and weak correlation coefficients), or even lead to wrong conclusion about vaccine efficacy. As example, an inversion of the sign of correlations was observed for intermediate VL upper-threshold without confirmation measure, between 3.5 and 4.8 log_10_ copies/mL in DALIA and between 4.0 and 4.5 log_10_ copies/mL in LIGHT. The use of a VL upper-threshold of at least 100 000 copies/mL with a confirmation measure allowed to retrieve at least 50% of the magnitude of the correlation between the setpoint and nAUC estimated on complete data (LIGHT: 50%, ILIADE: 94%, DALIA: 83%).

**Figure 7.**
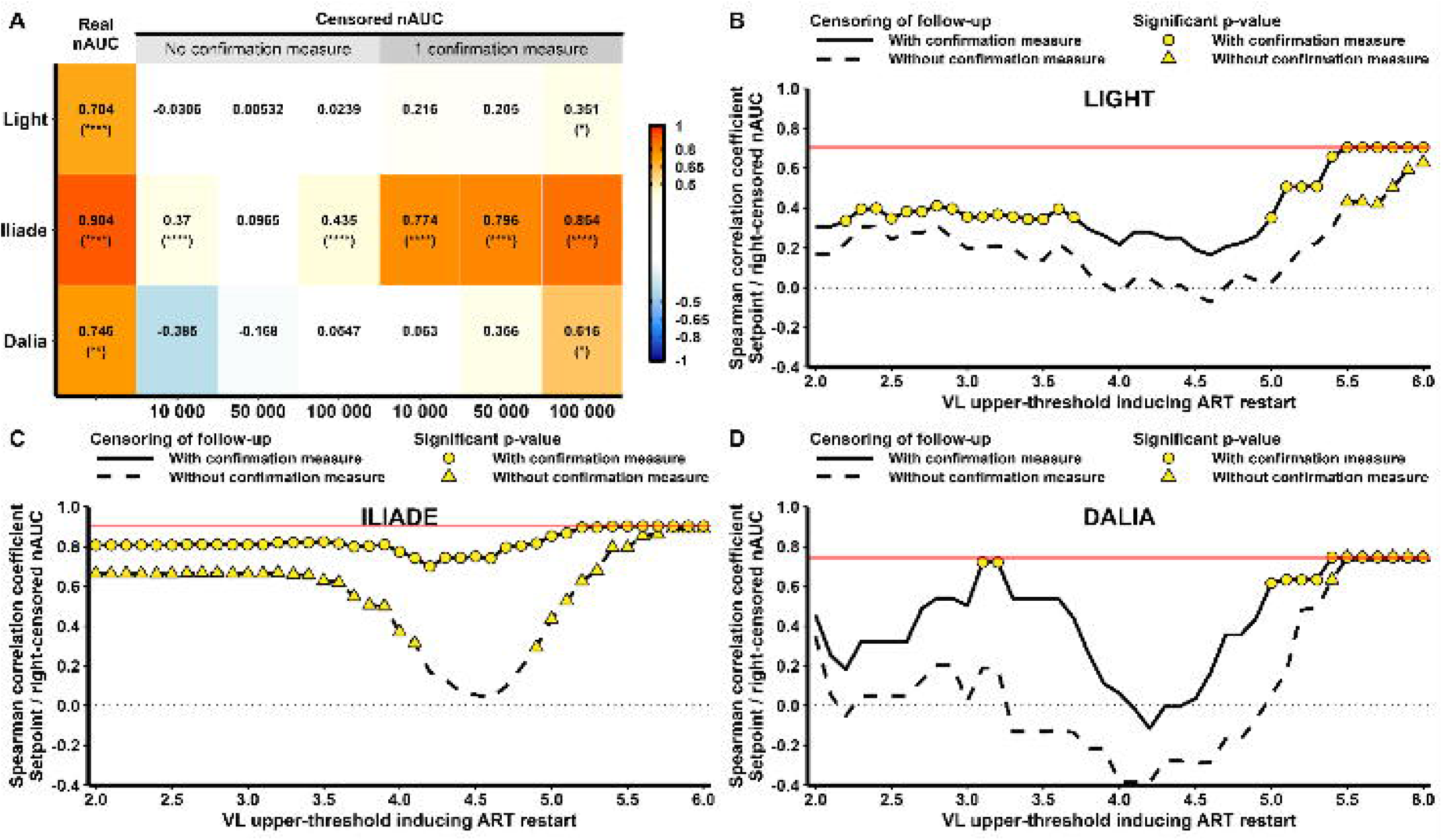
Spearman correlations between the setpoint and nAUC calculated on right-censored viral dynamics according to VL upper-thresholds. (A) Heatmap of Sperman correlations in each trial (y-axis) between the setpoint and nAUC, whether with or without censoring of follow-up, and with or without consideration of a confirmation measure before premature ART restart. Three commonly used VL upper-thresholds are considered: 10 000 cp/mL, 50 000 cp/mL and 100 000 cp/mL. Significant positive and negative correlations are highlighted in red and blue, respectively. **(B-D)** Analysis of Spearman correlations between the setpoint estimated on complete virological data, and the nAUC estimated on right-censored viral dynamics. The x-axis represents the VL upper-thresholds above which censoring of follow-up was considered. The y-axis represents the value of the correlation coefficient between the setpoint and the right-censored nAUC. Solid and dashed lines represent the results with or without a confirmation measure in the calculation of the right-censored nAUC, respectively. Yellow symbols (circles and triangles for with or without confirmation, respectively) highlight significant correlations (adjusted p-value ≤ 0.05). The horizontal gray dotted line represents the transition between positive and negative correlations. The horizontal red solid line represents the value of the correlation coefficient between the setpoint and the nAUC estimated on complete data. **(B)** LIGHT trial. **(C)** ILIADE trial. **(D)** DALIA trial.

## Discussion

This analysis of virological criteria in three different ATI clinical trials showed that the design of ATI phase and the definition of the virological endpoints are crucial for the evaluation of HIV cure strategies. When comparing the different virological criteria between the ATI trials, we found that especially the results of time-dependent virological criteria varied according to the design of ATI phase, i.e. ATI duration, sampling frequencies and intervals between viral load measurements. Our study identified the nAUC as the virological criteria the most strongly correlated with the setpoint. It has to be noted that, unlike the setpoint, the nAUC can be observed regardless of the design of ATI phase. Furthermore, we showed that the use of a too conservative threshold for virological ART re-start criterion can induce an increasing risk of missing important effects of the immune intervention on viral control. However, the use of a high value, i.e. 100 000 copies/mL reinforces the value of nAUC as a valid proxy of the setpoint. In particular, if the interest is in the post-replication control of infection, through the so-called setpoint studies (16), we showed that the nAUC is a valuable endpoint even in case of censored follow-up (28). Our results of correlations between virological criteria were consistent with those reported in other trials (17). The absence of correlation between time to rebound and setpoint has already been reported in other studies such as in the pooled analysis of 6 ACTG trials (19). Biologically, this could be explained by the fact that the initiation of the viral replication and the control of it afterwards are driven by different biological mechanisms. A recall T cell response stimulated by the start of the viral replication is potentially responsible for post-replication control (3,29). Hence, the pre-ART viral load is more correlated to the viral setpoint (19) than the time to rebound. Consequently, this absence of correlation emphases the necessity of choosing the most accurate and relevant primary endpoints that is in accordance with the objective of the trial. In the present work, it is also interesting to point out that the viral slope is also weakly correlated with the viral setpoint, suggesting that the secondary immune response is playing a bigger role on the control of viral replication after the peak as it is observed during primary infection.

This study is based on three clinical trials and thus the generalizability of our result may be limited. However, the information brought by real observed data from existing clinical trials helps to go beyond simulations (16) and the results were consistent with results reported in other additional trials (17,19). When the intervention and its expected impact is based on other mechanism than active immune control of viral replication such as passive immunization through neutralizing antibodies (Sneller et al. Science Trans Med 2019) then other endpoints and designs should be considered.

The statistical requirement that ART should not be re-initiated before confirmed viral load of more than 100 000 copies/mL is in agreement with what is recommended when the interest is in the viral setpoint (14). This does not preclude to consider all additional criteria (clinical and CD4 count) for patients’ safety. Specific statistical approaches have been developed to take into account incomplete information in such context (28).

Beyond statistical and clinical considerations, patient’s perception should be taken into account and recent studies have shown the good acceptability of ATI trials (12,13,30) as long as severe side effects are avoided (31). According to these studies, a part of the respondents would not accept any sustained period of viremia (35% in (32)). This would potentially restrict the inclusion rate in the trials. Also, the preferred frequency of biological measurements and clinical monitoring during ATI was monthly (32). Biotechnological improvements making measurements at home feasible could help coping with this patient’s preference (33).

### Key recommendations for future HIV therapeutic trials with ATI evaluating the ability of a treatment to control viral replication in the absence of ART

Results provided by this work allowed us to make some recommendations. First, the time-averaged area under the viral rebound curve during the ATI phase (nAUC here) has been shown to be the optimal virological criterion to use as proxy to the setpoint and consequently as primary virological endpoint. Secondly, the optimal design of ATI phase allowing a good balance between minimization of the risk for patients and the maximization of the capacity to evaluate the effect of intervention on viral dynamics includes (1) a duration of the ATI phase of more than 12 weeks and no longer than 24 weeks to ensure patients safety, (2) regular monitoring during the early phase of the viral rebound dynamics, with viral load measurements every 1 to 2 weeks until reaching the setpoint (i.e., around week 12), and (3) viral load measurements every 2 to 4 weeks thereafter. Finally, the use of the VL upper-threshold of 100 000 copies/mL as virological ART re-start criteria appears as the best trade-off limit the risk for patients and capture enough information to keep nAUC as a good proxy of the setpoint. These results and proposals could help to optimize the design of clinical trials testing HIV cure strategies.

## Contributions

Y.L., M.P., and R.T. conceptualized the study. M.A. accessed and verified data, and conducted the formal analysis. M.P. and R.T. supervised the analysis. All authors contributed to data interpretation. M.A created the first draft of the figures, and M.A. and R.T. wrote the first draft of the manuscript. All authors critically revised and edited the manuscript and approved the final version for submission.

## Data availability

The data that support the findings of this study are available from the corresponding author, R.T, upon reasonable request.

