## Supplementary Figures for "Definition of viroIogical endpoints improving the design of HIV cure strategies using analytical antiretroviral treatment interruption"

**Figure S1 –** **Flow charts describing the three therapeutic vaccine trials. (A)** LIGHT trial. **(B)** DALIA trial. **(C)** ILIADE trial.

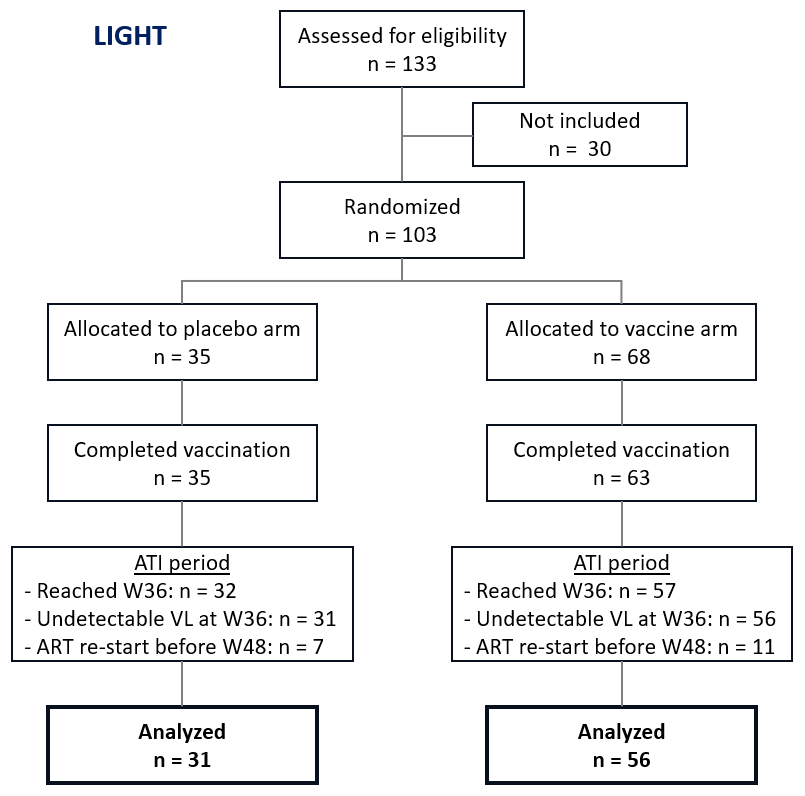

**A**

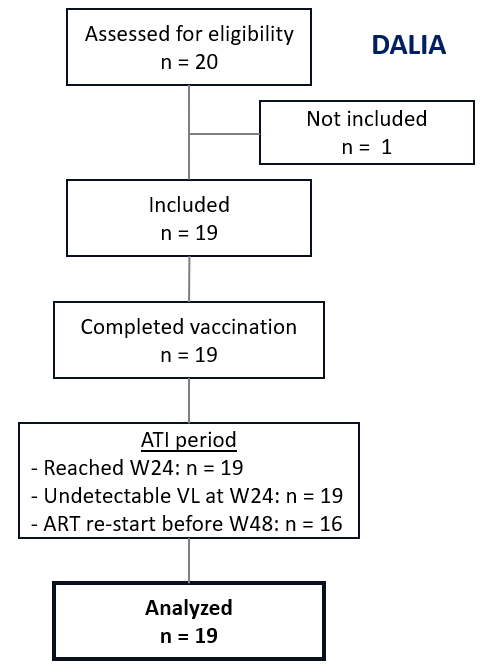

**B**

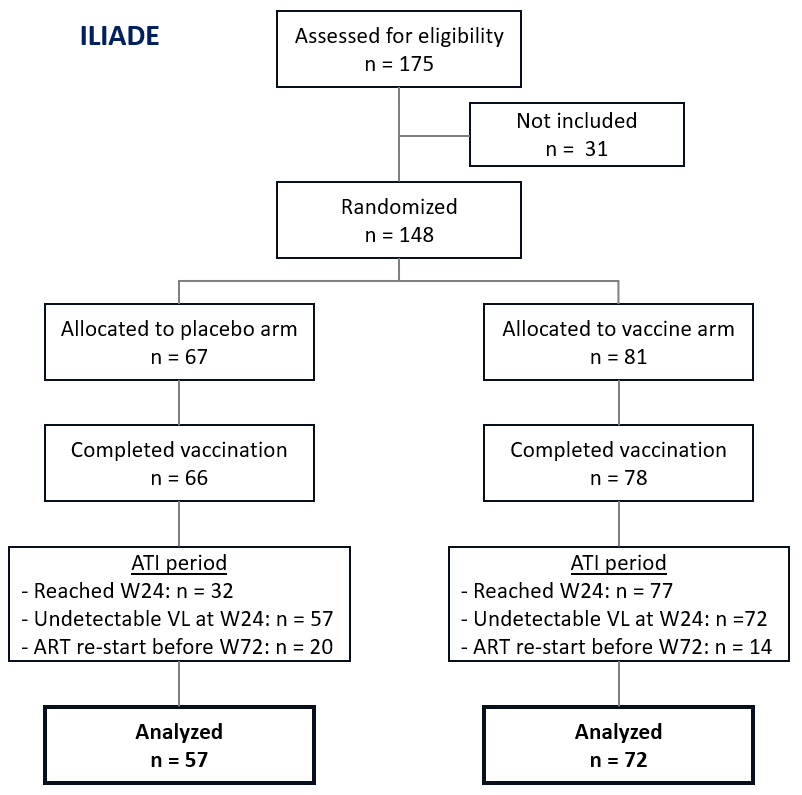

**C**

**Figure S2 –** **The proportions of patients during ATI phase with viral load within specific ranges of VL values.** The percentages are provided for each trial. Only percentages higher than 5% are displayed in bars.

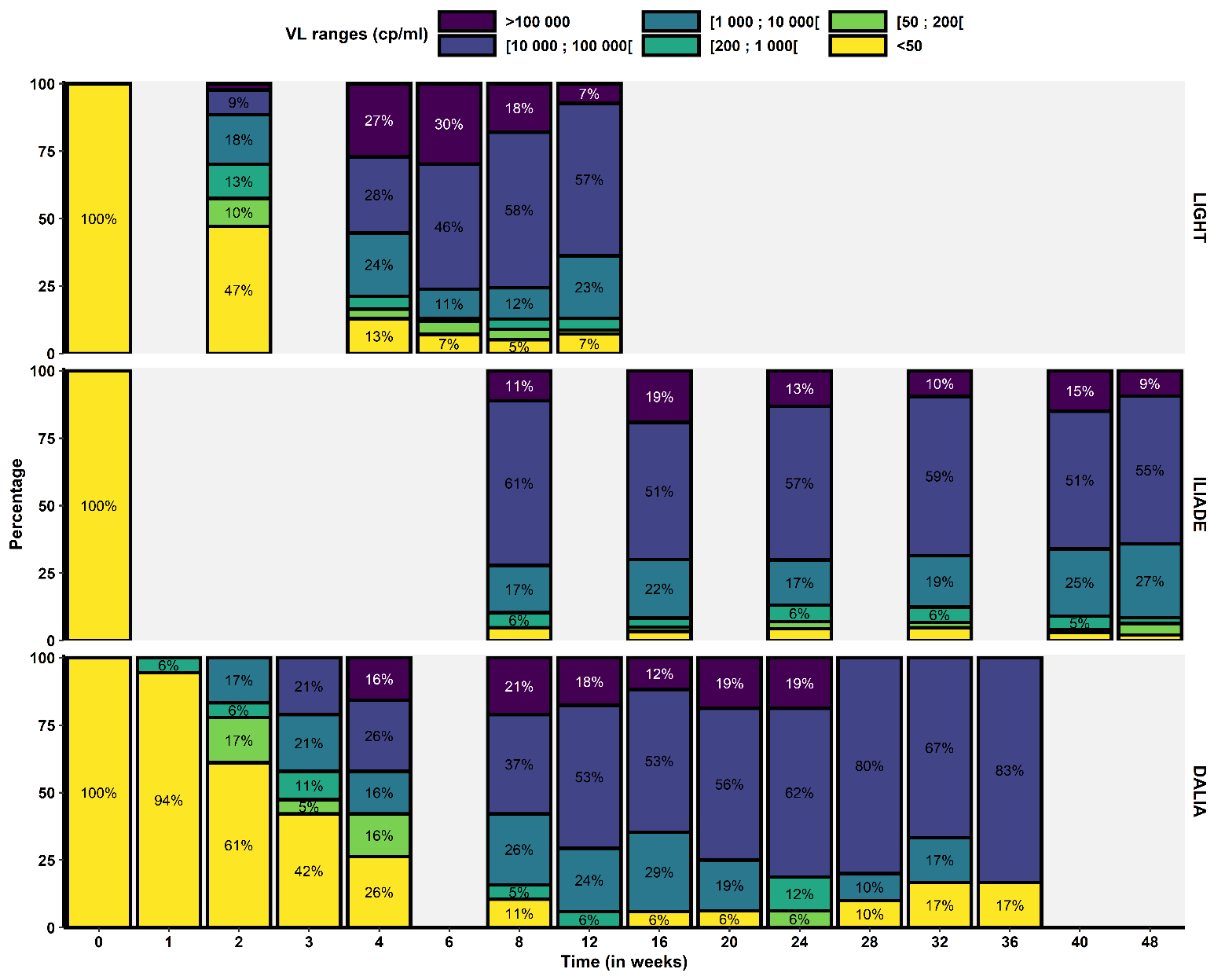

**Figure S3 – Example of the impact of study design on the value of the VL peak during ATI phase.** Thick black line represents an example of real dynamic of viral rebound during ATI, simulated with a mathematical model. Thin colored lines and colored symbols represent observed dynamics extracted from the black curve according to study design: blue line and blue circles for observations collected every 2 weeks (i.e., LIGHT study design), and orange line and orange triangles for observations collected every 8 weeks (i.e., ILIADE study design). Filled circles and triangles represent the peak of the observed dynamics extracted with LIGHT and ILIADE study designs, repsepectively.

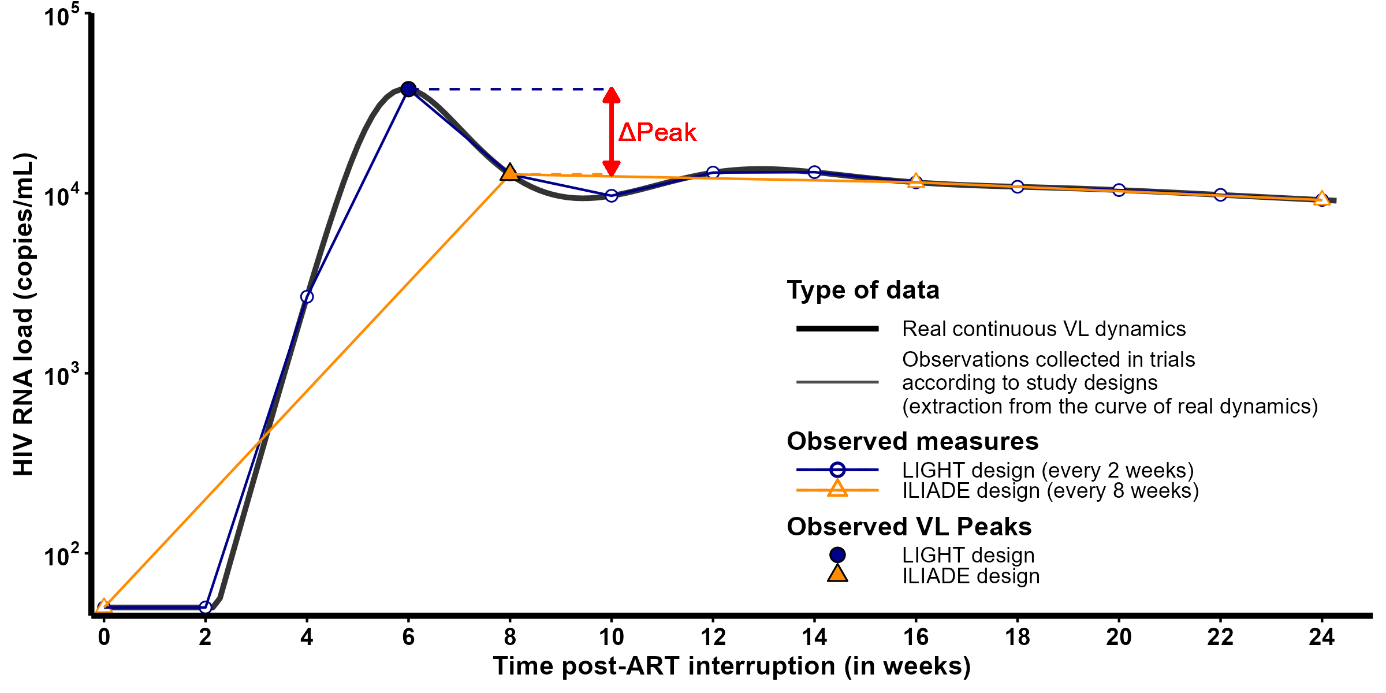

**Figure S4 – Heatmap of pairwise Spearman correlations between the eleven virological criteria studied in the analysis, in the three clinical studies. (A)** LIGHT clinical trial (n=87). **(B)** ILIADE clinical trial (n=129). **(C)**  DALIA clinical trial (n=19). **(A-C)** Correlations higher than 0.5 are colored in orange-red palette and correlations lower than -0.5 in blue palette. P-values adjusted for multiplicity testing are indicated in brackets: *: p ≤ 0.05, **: p ≤ 0.01, ***: p ≤ 0.001, ****: p ≤ 0.0001. Abbreviations: nAUC, time-averaged area under the curve; TTR: time to rebound.

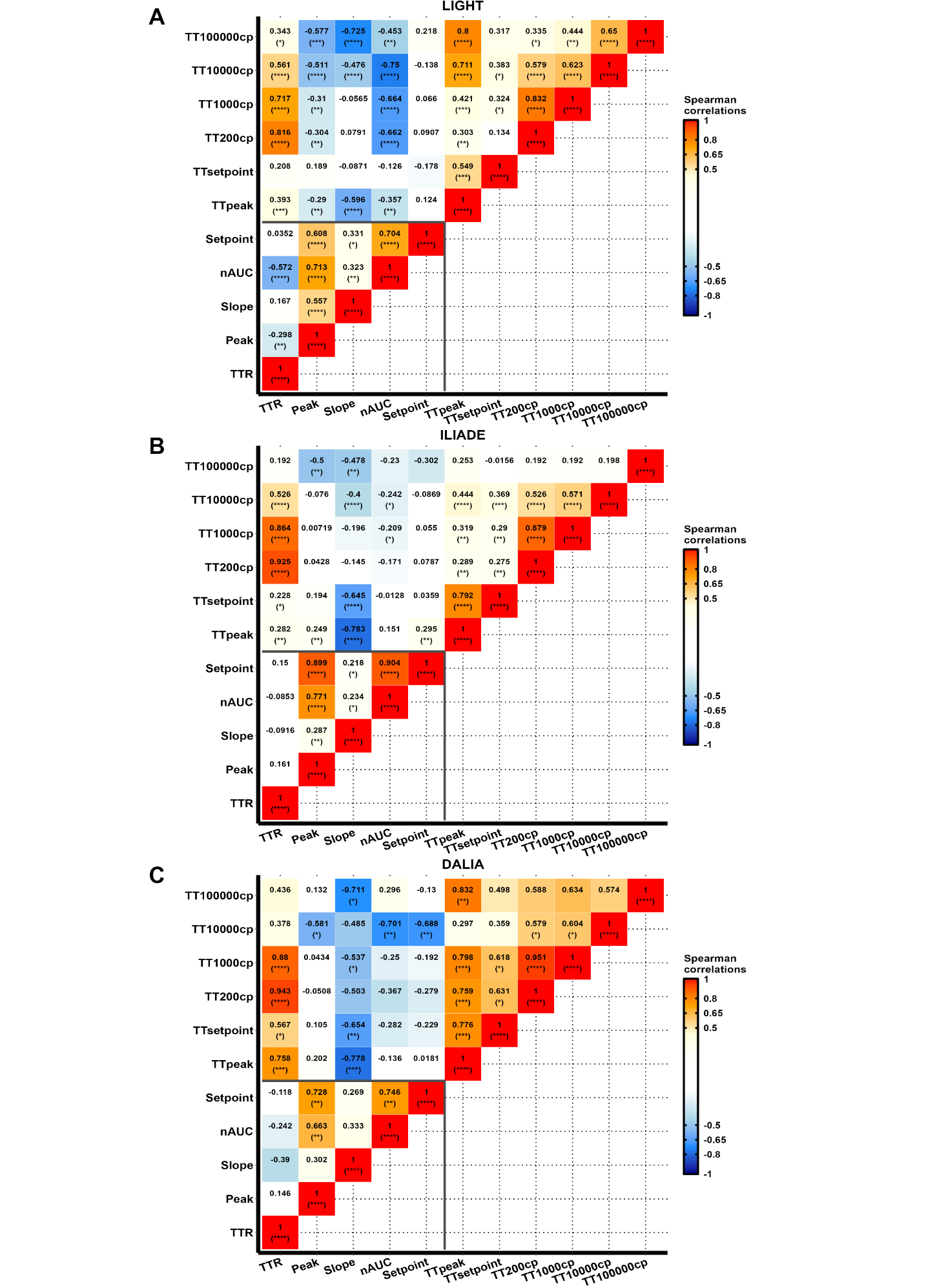

**Figure S5 – Pearson correlations matrices and scatter plots between the five main virological criteria observed in the three clinical trials. (A)** LIGHT clinical trial (n=87). **(B)** ILIADE clinical trial (n=129). **(C)** DALIA clinical trial (n=19). **(A-C)** The upper panels represent pairwise Pearson’s correlation coefficients, with positive correlations colored in red, negative correlations in blue, and P values indicated in brackets. The lower panels represent scatter plots with solid red lines corresponding to linear trend lines and shaded areas to confidence intervals. The middle panels represent the distributions of endpoints. P-values adjusted for multiplicity testing are indicated in brackets: *: p ≤ 0.05, **: p ≤ 0.01, ***: p ≤ 0.001, ****: p ≤ 0.0001. Abbreviations: nAUC, time-averaged area under the curve; TTR: time to rebound.

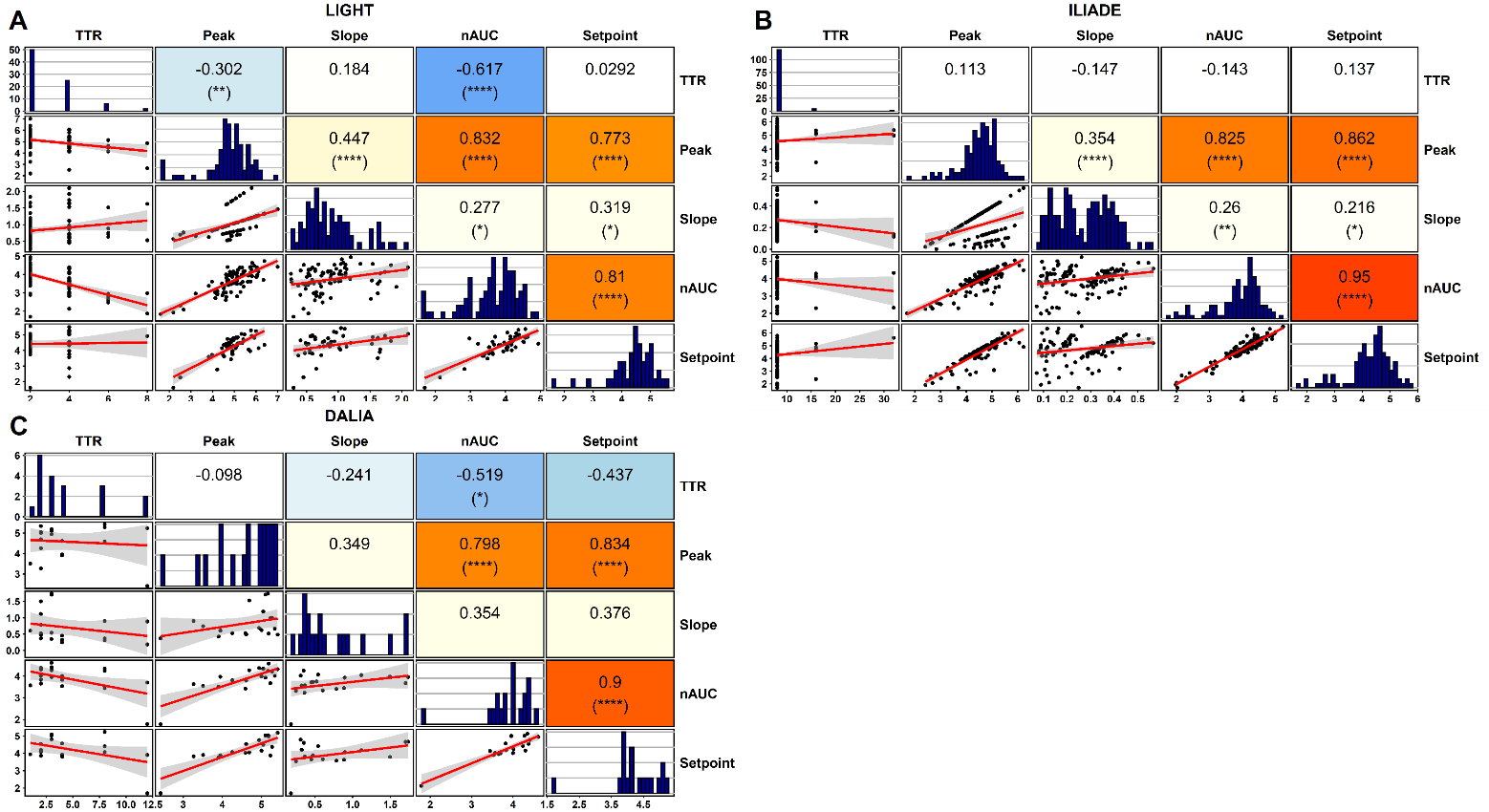
