## Supplementary materials for "Definition of viroIogical endpoints improving the design of HIV cure strategies using analytical antiretroviral treatment interruption"

**Table S1** – **Virological measurements (in log_10_ scale) collected in the three HIV therapeutic vaccine trials during ATI phase, and analyzed in the study.** Blue areas highlight time of measurements beyond time of evaluation of primary virological endpoint (i.e., long-term follow-up).

| Weeks during ATI phase | | **LIGHT** (n=87) | | **ILIADE** (n=129) | | **DALIA** (n=19) |
| --- | --- | --- | --- | --- | --- | --- |
|  |  | Placebo (n=31) | Vaccine (n=56) | Control (n=57) | IL2 (n=72) |  |
| W0 | Number pat. | 31 | 56 | 57 | 72 | 19 |
|  | Censored (%) | 31 (100) | 56 (100) | 57 (100) | 72 (100) | 19 (100) |
|  | Mean [IQR] | 1.60 [1.60 ; 1.60] | 1.60 [1.60 ; 1.60] | 1.70 [1.70 ; 1.70] | 1.70 [1.70 ; 1.70] | 1.70 [1.70 ; 1.70] |
| W1 | Number pat. |  |  |  |  | 18 |
|  | Censored (%) |  |  |  |  | 17 (94) |
|  | Mean [IQR] |  |  |  |  | 1.75 [1.70 ; 1.70] |
| W2 | Number pat. | 31 | 56 |  |  | 18 |
|  | Censored (%) | 14 (45) | 23 (41) |  |  | 11 (61) |
|  | Mean [IQR] | 2.48 [1.60 ; 3.13] | 2.49 [1.60 ; 3.32] |  |  | 2.05 [1.70 ; 2.13] |
|  | P-value^(1)^ | P = 0.80 | |  |  |  |
| W3 | Number pat. |  |  |  |  | 19 |
|  | Censored (%) |  |  |  |  | 8 (42) |
|  | Mean [IQR] |  |  |  |  | 2.79 [1.70 ; 3.39] |
| W4 | Number pat. | 31 | 54 |  |  | 19 |
|  | Censored (%) | 3 (10) | 8 (15) |  |  | 5 (26) |
|  | Mean [IQR] | 4.18 [3.47 ; 5.41] | 4.09 [3.42 ; 4.95] |  |  | 3.26 [1.72 ; 4.51] |
|  | P-value^(1)^ | P = 0.68 | |  |  |  |
| W6 | Number pat. | 30 | 54 |  |  |  |
|  | Censored (%) | 2 (7) | 4 (7) |  |  |  |
|  | Mean [IQR] | 4.41 [4.18 ; 4.95] | 4.32 [4.04 ; 5.13] |  |  |  |
|  | P-value^(1)^ | P = 0.80 | |  |  |  |
| W8 | Number pat. | 29 | 49 | 55 | 71 | 19 |
|  | Censored (%) | 1 (3) | 3 (6) | 2 (4) | 4 (6) | 2 (11) |
|  | Mean [IQR] | 4.16 [3.74 ; 4.76] | 4.29 [4.15 ; 4.90] | 4.30 [4.06 ; 4.77] | 4.20 [3.86 ; 4.80] | 4.15 [3.90 ; 5.00] |
|  | P-value^(1)^ | P = 0.43 | | P = 0.53 | |  |
| W12 | Number pat. | 24 | 45 |  |  | 17 |
|  | Censored (%) | 1 (4) | 3 (7) |  |  | 0 (0) |
|  | Mean [IQR] | 4.01 [3.78 ; 4.62] | 4.06 [3.74 ; 4.67] |  |  | 4.36 [3.96 ; 4.66] |
|  | P-value^(1)^ | P = 0.74 | |  |  |  |
| W16 | Number pat. |  |  | 53 | 67 | 17 |
|  | Censored (%) |  |  | 2 (4) | 2 (3) | 1 (6) |
|  | Mean [IQR] |  |  | 4.26 [3.97 ; 4.93] | 4.22 [3.88 ; 4.75] | 4.27 [3.91 ; 4.79] |
|  | P-value^(1)^ |  |  | P = 0.59 | |  |
| W20 | Number pat. |  |  |  |  | 16 |
|  | Censored (%) |  |  |  |  | 1 (6) |
|  | Mean [IQR] |  |  |  |  | 4.31 [4.01 ; 4.76] |
| W24 | Number pat. |  |  | 49 | 65 | 16 |
|  | Censored (%) |  |  | 2 (4) | 3 (5) | 0 (0) |
|  | Mean [IQR] |  |  | 4.22 [3.97 ; 4.93] | 4.16 [3.88 ; 4.67] | 4.15 [4.13 ; 4.75] |
|  | P-value^(1)^ |  |  | P = 0.49 | |  |
| W28 | Number pat. |  |  |  |  | 10 |
|  | Censored (%) |  |  |  |  | 1 (10) |
|  | Mean [IQR] |  |  |  |  | 4.04 [4.14 ; 4.64] |
| W32 | Number pat. |  |  | 43 | 62 | 6 |
|  | Censored (%) |  |  | 2 (5) | 3 (5) | 1 (17) |
|  | Mean [IQR] |  |  | 4.13 [3.90 ; 4.70] | 4.19 [3.94 ; 4.72] | 3.97 [3.95 ; 4.66] |
|  | P-value^(1)^ |  |  | P = 0.72 | |  |
| W36 | Number pat. |  |  |  |  | 6 |
|  | Censored (%) |  |  |  |  | 1 (17) |
|  | Mean [IQR] |  |  |  |  | 4.01 [4.06 ; 4.67] |
| W40 | Number pat. |  |  | 40 | 60 |  |
|  | Censored (%) |  |  | 2 (5) | 1 (2) |  |
|  | Mean [IQR] |  |  | 4.23 [3.95 ; 4.77] | 4.16 [3.76 ; 4.73] |  |
|  | P-value^(1)^ |  |  | P = 0.53 | |  |
| Week 48 | Number pat. |  |  | 37 | 58 |  |
|  | Censored (%) |  |  | 1 (3) | 1 (2) |  |
|  | Mean [IQR] |  |  | 4.19 [3.86 ; 4.81] | 4.14 [3.72 ; 4.65] |  |
|  | P-value^(1)^ |  |  | P = 0.74 | |  |

ART: Antiretroviral treatment, ATI: Analytical treatment interruption, IQR: Interquartile range, Pat.: Patient.

^(1)^ P-values of pairwise comparisons (Mann-Whitney U test) in the different trials between the therapeutic and the control groups.
